# Assessing the Impact of Non-exhaust Emissions on the Asthmatic Airway (IONA) Protocol for a randomised three exposure crossover study

**DOI:** 10.1101/2024.01.30.24301985

**Authors:** James Scales, Hajar Hajmohammadi, Max Priestman, Luke C. McIlvenna, Ingrid E. de Boer, Haneen Hassan, Anja H. Tremper, Gang Chen, Helen E. Wood, David C. Green, Klea Katsouyanni, Ian S. Mudway, Christopher Griffiths

## Abstract

**Background:** People living with asthma are disproportionately affected by air pollution, with increased symptoms, medication usage, hospital admissions and the risk of death. To date there has been a focus on exhaust emissions, but traffic-related air pollution (TRAP) can also arise from the mechanical abrasion of tyres, brakes, and road surfaces. Non-exhaust emissions (NEE) currently make up a greater proportion of TRAP by mass than exhaust emissions. With the increasing weight of vehicle fleets due to electrification, and increasing uptake of larger vehicles, it is likely that NEE will continue to be an increasing health challenge.

These NEE remain unregulated and underexplored in terms of their health impacts, particularly in vulnerable groups such as people living with asthma. To date, few real-world studies have attempted to explore the impacts of non-exhaust emissions on human health. We therefore created a study with the aim of investigating the acute impacts of NEE on the lung function and airway immune status of asthmatic adults.

**Methods:** The IONA study will expose adults with asthma in random order at three locations in London selected to provide the greatest contrast in the NEE components within TRAP. Health responses will be assessed before and after each exposure, with lung function measured by spirometry as the primary outcome variable.

**Discussion:** Collectively this study will provide us with valuable information on the health effects of NEE components within ambient PM_2.5_ and PM_10_, whilst establishing a biological mechanism to help contextualise current epidemiological observations.

**Trial registration:** IRAS Number 320784 at https://clinicaltrials.gov/

## Introduction

The global burden of disease survey estimated that air pollution contributed to one in eight deaths in 2019 (Murray et al, 2019). These effects are largely attributed to long-term exposures to fine particulate matter, PM_2.5_ (particulate matter with an aerodynamic diameter on average less than 2.5 μm in diameter) and nitrogen dioxide (NO_2_), but short-term exposures are also associated with significant health impacts in vulnerable groups, such as people with asthma, including worsening of symptoms (Su et al, 2023), increased hospitalization (Delamater et al, 2012)(Ashworth et al, 2021) and death (Liu et al, 2019).

Traffic as a source of PM (particulate matter) has received considerable attention (Health Effects Institute, 2022), with evidence that distance to roads, traffic density/composition (Boogaard et al, 2022), and exhaust emission tracers such as elemental/black carbon, particle number concentration and NO_2_ (Janssen, et al 2011) are all associated with both acute and long-term negative health effects. There is therefore a need to better understand the contribution of different components of TRAP to these adverse effects, to provide both better advise to vulnerable groups and help shape government policy to reduce all traffic emissions. To date the contribution of traffic-derived air pollution research has focused on the contribution of exhaust emissions, whilst particulates from NEE have been understudied (Fussell et al, 2021). Particles arising from tyre and brake wear, as well as the resuspension of road dust, represent a greater proportion of roadside PM by mass than direct exhaust emissions (Piscitello et al, 2021). As exhaust emissions decrease as nations strive to meet their NetZero commitments, greater attention on non-exhaust sources is urgently required to evaluate their relative hazard compared with other pollutant sources (AQEG, 2019). NEE remain unregulated, and their health impacts under-explored, particularly in vulnerable groups. Air pollution has been shown to significantly impact people living with asthma, increasing prevalence of childhood asthma (Khreis et al, 2017), exacerbations in adults (Orellano et al, 2017) and hospital admissions (Zheng et al, 2015). As such, research into the health effects of NEE on people with asthma is urgently needed.

Disentangling the relative contributions of different chemical mixtures within ambient PM has proven challenging in both short- and long-term epidemiological studies, due to the high degree of correlation between components with a common source. Currently there is no firm consensus as to which components of PM_2.5_ or PM_10_ (particulate matter with an aerodynamic diameter on average less than 10 μm in diameter) present the greatest hazard to the population (Lippmann et al, 2013). Previous work (Steenhof et al., 2014, Strak et al., 2013b, Steenhof et al., 2013, Strak et al., 2013a, Strak et al., 2012, Steenhof et al., 2011, Strak et al., 2011) has demonstrated the feasibility of tackling this challenge using a quasi-experimental human real-world exposure crossover design in healthy subjects. Whilst this work showed the key role played by primary gaseous and particulate emissions, the study focused on responses in healthy adults and did not examine biological pathways that lead to symptom exacerbation in vulnerable populations such as in individuals with asthma. These studies examined a range of metals such as iron (Fe), copper (Cu), nickel (Ni) and vanadium (V), reflecting their capacity to cause oxidative stress, but did not explicitly relate these to NEEs, nor employ them for detailed source apportionment.

To date, whether real-world NEE are causally related to worsening of asthma has not been widely studied. Therefore, the primary aim of this study is to assess and compare the acute changes in lung function and airway inflammation in asthmatics when exposed to three microenvironments with contrasting non-exhaust traffic-related air quality profiles. The contrasting traffic profiles increase the contrast in (?) PM_10_ and PM_2.5_ from brake wear, tyre wear, and road abrasion, although tyre and road wear are often combined into a single source tyre and road wear particles (TRWP).

## Methods and analysis

### Study design

The IONA study will employ a randomised three condition crossover panel design. We will recruit 48 adults (aged over 18 years) residing in London, with mild-to-moderate allergic asthma that started in childhood.

### Inclusion criteria

1. Doctor-diagnosed asthma starting on or before age 16 years
2. Prescribed regular inhaled corticosteroid medication
3. Able to use a static exercise bicycle for the study duration

### Exclusion criteria

1. Tobacco smoking, or living with smoker
2. BMI > 30
3. Asthma hospitalisation within 12 months
4. Three or more asthma episodes requiring oral corticosteroid medication within 12 months
5. Other major lung disease
6. Chest surgery within 6 months
7. Unable to give informed consent
8. Occupational exposure to PM or high levels of air pollution^**a**^
9. Under the age of 18
10. Individuals at any stage of pregnancy
11. Currently Breast-feeding

^**a**^ For the purposes of this study occupational exposure is defined as people who work as taxi drivers, couriers, waste removal drivers and utility services drivers. If other occupations with potentially high exposure to air pollution approach the study. The study team will discuss eligibility of the occupation within the project management group, comprising of experts in air pollution and asthma management.

### Aims/outcomes

#### Primary

- To compare the acute health effects (changes in lung function and airway inflammation) in mild/moderate asthmatics of exposure to PM_2.5_ and PM_10_ at three selected microenvironments with contrasting contributions from brake wear and tyre and road wear (TRWP).

#### Secondary

- Provision of an air pollutant database (PM_2.5_ and PM_10_ mass and chemical composition, PNC and NO_2_) and a time series of source-apportioned PM_2.5_ and PM_10_ covering all exposure days at the three selected sites.
- To examine the relationship between variations in daily non-exhaust source fractions derived from brake and TRWP, with the deposited dose, based on metal/metalloid concentrations determined in the nasal airways, relating these measures of biological dose to the physiologic and immunologic responses observed.
- To establish a biobank of plasma, nasal lavage, and urine samples for future work examining molecular signatures of exposure and response to NEE.

#### Outcome measures

- Primary: Lung function as measured by Forced Expiratory Volume in one second (FEV_1_), this measure has been selected as the primary outcome based upon previous work showing detrimental effects of time spent in an urban atmosphere on lung function (McCreanor et al, 2007).
- Secondary: Spirometry (FVC, FVC/FEV_1_ ratio), Fractional Expired Nitric Oxide (FeNO), oscillometry (R5, R20, T5, T20, AX), asthma symptoms, nasal mucosal immune responses.
- Biobanks: urine and blood plasma samples will be collected for future analysis.

### Study interventions

Due to the complexity of multipollutant aerosol mixtures within exhaust and NEE, we will utilise an efficient randomised cross-over semi-experimental design investigating short-term respiratory health impacts in non-smoking adults with mild-moderate asthma during and after sequential standardised exercise exposures to three contrasting air quality environments, comprising:

1. A busy road junction characterized by stop-go traffic to enhance emissions from brake wear
2. High speed continuous traffic, to enhance TRWP emissions
3. An urban background location away from nearby traffic sources

Sites have been selected to provide the greatest contrast in emission sources and to be a cost-effective method of establishing the relationship between PM source contributions and acute health effects within the real-world context.

The overall protocol, and the participant flow through the three separate exposures, are summarised in Figure 1. Following consent and initial assessment, participants will visit the three field-testing sites in random order. During the exposure protocol participants will ride on a static bike for 15 minutes at time, with 15-minute rest periods, for a total of six cycling periods at a standardised intermittent intensity. Participants will perform identical respiratory health assessments preceding, halfway through, and following the exercise task. In parallel to exposures at sites, we will perform real-time measurements of PM_10_ and PM_2.5_, PNC and gaseous pollutants ozone (O3), NO_2_, black carbon. The protocol for the three exposure visits will be identical and each visit will be separated by a minimum two-week washout period, with testing taking place between May and October 2023.

**Figure 1:**
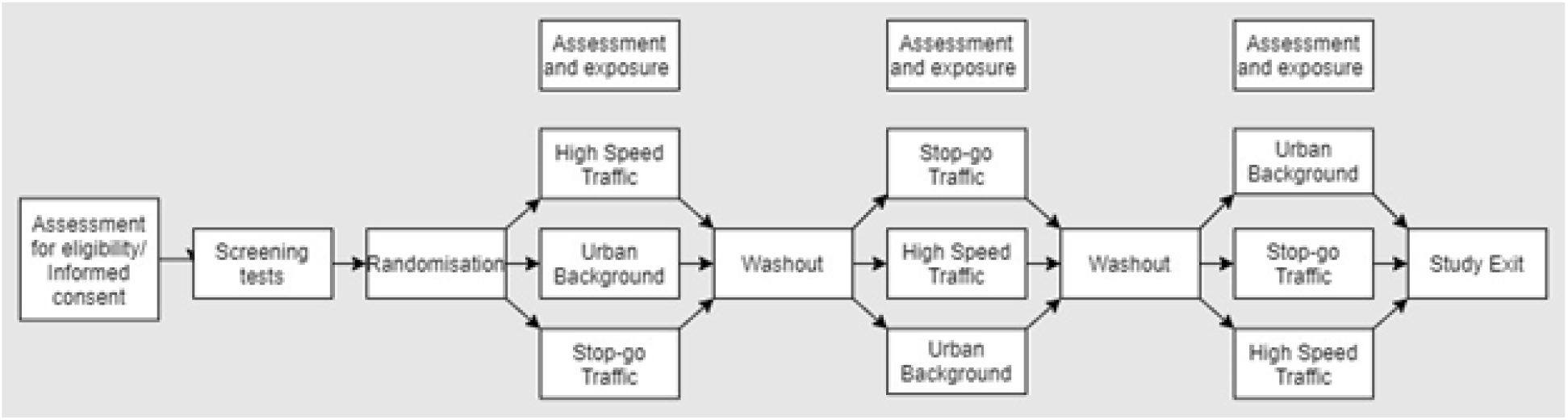
Diagram of participant flow through IONA study.

To control for variations in air quality, exposure will occur between 11:00am and 13:30pm on weekdays. Between pre- and post-health assessments, participants will be asked to cycle on a static exercise bike at a watt equivalent to 60% of estimated heart rate. This exercise intensity was chosen to maximise inhaled doses of PM and gaseous pollutants over the exposure period while remaining below the first ventilation threshold to reduce variation in ventilatory drift, and minimising discomfort for the participants. Physiological drift will be monitored by Ratings of Perceived Exertion questionnaire and real-time monitoring of the participant’s heart rate.

### Exposure site selection

Three sites will be used for the exposure visits. The two air quality supersites in London, located in a suburban park in South London (Honor Oak Park) and close to a major trunk road in Central London (Marylebone Road), will be used as a basis for this study. These sites are equipped with the ability to collect time-resolved chemical composition measurements to quantify the non-exhaust component of PM. An additional measurement location will be established below a high-speed flyover (A40 flyover) using a mobile measurement facility equipped with the same highly time-resolved chemical composition measurement capability as the supersites. This location provides the contrast in non-exhaust contributions, being next to the high speed A40 flyover.(Figure 2).

**Figure 2:**
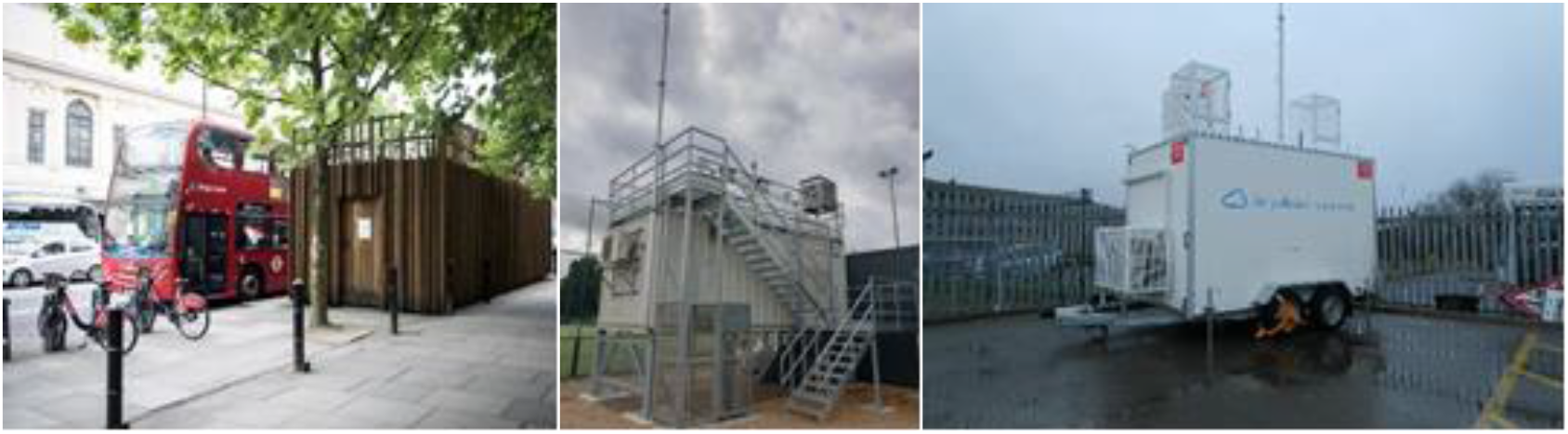
Air quality supersites: Marylebone Road (left), Honor Oak Park (centre), mobile measurement station (not in location).

The Marylebone Road station is located 2m from kerbside, this major trunk road carries 51,000 vehicles per day - 4% Heavy goods vehicles (HGV), 3% Buses, 16% light goods vehicles (LGV); (Department for Transport, 2022) the average speed is 34 kph (21 mph). The stop-go traffic on this congested road increases brake wear relative to tyre wear and resuspension. This has been confirmed by the London Atmospheric Emissions Inventory, which estimates 51% of NEE as brake wear compared to 39% as resuspension and 10% as tyre wear.

The proposed location on the A40 flyover is 5m from the kerbside of the busy A40 flyover, carrying 93,000 vehicles per day - 6% HGVs, 17% LGVs; (Department for Transport, 2022) average speed was 33-59 kph (21-37mph). This road is a major route into and out of Central London, the high number of HGV and associated vehicle weight will lead to increased resuspension and tyre wear. (Fussell et al, 2022; USEPA, 2011.) Due to the higher speeds and more free-flowing traffic, the London Atmospheric Emissions Inventory, estimates this location to have only 85% of the brake wear emissions of the Marylebone Road measurement location thereby increasing the contrast between the two traffic locations.

### Sample size

The study uses an efficient Williams design (Williams, 1949) to achieve a significance level of 0.05, a Δ (Difference between the reduction in FEV_1_ between two sites) of 4.69 with a standard deviation of 6.4 (McCreanor et al., 2007). At 91.5% power, 30 participants are required. To account for a dropout rate of 30%, and uncertainty regarding exact exposures on study days, we intend to recruit 48 participants in total. Since we will randomise the participants to a sequence for their exposures to the three sites, we will have six sequences (ABC/ BCA/ CBA/ ACB/ BAC/ CAB). We will define 12 groups of up to 6 individuals each and then assign a group to each participant using randomly numbered sheets sealed in opaque envelopes.

### Recruitment

#### Primary Care

A Participant Identification Centre site approach will be used to recruit participants. Initially, participants will be recruited from general practice offices across Central and East London. Eligible participants identified by searches will be approached with an SMS invitation to join the study via secure electronic mail.

#### Non primary care

Participants will also be recruited through other avenues. We will target people with asthma who study or work at Queen Mary, University of London and Imperial College London. Posters, lecture visits, and mailing lists will be used. Finally, we will recruit participants from the public through social media and posters.

#### Patient and Public Involvement

This study was developed with input from the Asthma and Lung UK Centre for Applied Research (AUKCAR) Public and Patient Involvement (PPI) group. The group contributed to the study by providing feedback on the study design and methods. As the study progresses, the PPI group will be invited to project management group meetings as advisors and partners to discuss analysis, and results and dissemination.

#### Preliminary assessment

Prior to assessment at exposure sites, participants will perform an exercise test to calculate exercise intensity (described below), perform post-bronchodilator spirometry and complete an Asthma Control Test (Nathan et al, 2004) to confirm asthma status. Spirometry assessment is defined in the data collection and analysis section below.

#### Estimated Vo_2max_ fitness assessment

To support standardisation of exercise intensity at exposure sites, participants will perform an estimated VO_2max_ test during initial assessment. Participants will be asked to perform an incremental exercise test following YMCA cycle ergometry protocols defined by The American College for Sports Medicine (Lippincott Williams and Wilkins, 2013). Results will be used to calculate estimated Vo_2max_ which will in turn will be used to calculate a target watt equivalent for 60% estimated Vo_2max_.

#### Exposure

To control for variations in air pollution, exposures will be in groups of between 2 and 6 participants at 11am until 13:30pm on weekdays. Participants will exercise on cycle ergometers (Wattbike Gen II, Wattbike Ltd, Nottingham, UK) at corresponding power output of 60% of their estimated Vo_2max_ for six 15-minute exercise bouts interspersed with 15-minute rest periods. This intensity is chosen to minimise cardiovascular drift and maximise inhaled dose. 15-minute rest periods are chosen to minimise cardiovascular drift, allow comparison to previous laboratory-based work and to facilitate multiple participants at the testing site.

#### Data collection before and after exposure

Before and after each exposure a battery of physiological measurements and samples will be taken. The timepoints of each measurement or sample are presented in an overview in table 2.

**Table 2:** Flow of participants through IONA study days.

|  | IONA: Schedule of assessment and exposure |  |  |  |  |  |  |  |
| --- | --- | --- | --- | --- | --- | --- | --- | --- |
|  | Pre-Study | Initial assessment | 3-day pre | Pre | Exposure | Post | 24-hour post | 3-days post |
|  |  | Phys lab | Self-assess | Field lab | Field lab | Field lab | Phys lab | Self-assess |
| Consent | x |  |  |  |  |  |  |  |
| Exercise test |  | x |  |  |  |  |  |  |
| FeNO |  | x |  | x | x | x | x |  |
| Oscillometry |  | x |  | x | x | x | x |  |
| Spirometry |  | x |  | x | x | x | x |  |
| Asthma control test |  | x |  |  |  |  |  |  |
| Asthma quality of life test |  | x |  |  |  |  |  |  |
| Asthma symptoms |  |  | x |  |  |  | x | x |
| Venepuncture |  |  |  | x |  |  | x |  |
| Nasal lavage |  |  |  | x |  | x | x |  |
| Urine Sample |  |  |  | x |  |  | x |  |
| Heart rate |  |  |  |  | x |  |  |  |

**Table 3:**
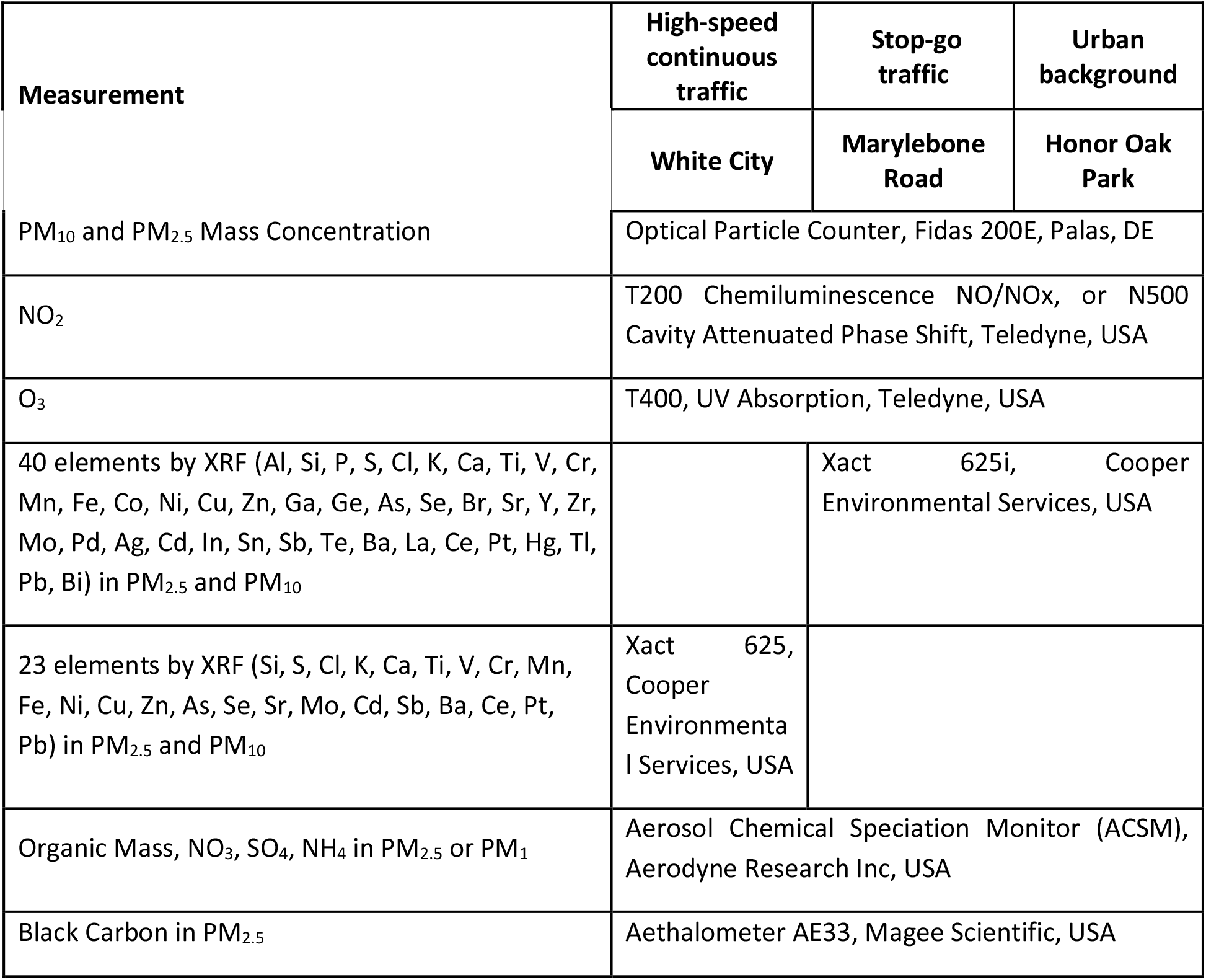
Air quantity station measurement configuration.

| Measurement | High-speed continuous traffic | Stop-go traffic | Urban background |
| --- | --- | --- | --- |
|  | White City | Marylebone Road | Honor Oak Park |
| PM<sub>10</sub> and PM<sub>2.5</sub> Mass Concentration | Optical Particle Counter, Fidas 200E, Palas, DE |  |  |
| NO<sub>2</sub> | T200 Chemiluminescence NO/NO<sub>x</sub>, or N500 Cavity Attenuated Phase Shift, Teledyne, USA |  |  |
| O<sub>3</sub> | T400, UV Absorption, Teledyne, USA |  |  |
| 40 elements by XRF (Al, Si, P, S, Cl, K, Ca, Ti, V, Cr, Mn, Fe, Co, Ni, Cu, Zn, Ga, Ge, As, Se, Br, Sr, Y, Zr, Mo, Pd, Ag, Cd, In, Sn, Sb, Te, Ba, La, Ce, Pt, Hg, Tl, Pb, Bi) in PM<sub>2.5</sub> and PM<sub>10</sub> |  | Xact 625i, Cooper Environmental Services, USA |  |
| 23 elements by XRF (Si, S, Cl, K, Ca, Ti, V, Cr, Mn, Fe, Ni, Cu, Zn, As, Se, Sr, Mo, Cd, Sb, Ba, Ce, Pt, Pb) in PM<sub>2.5</sub> and PM<sub>10</sub> | Xact 625, Cooper Environmental Services, USA |  |  |
| Organic Mass, NO<sub>3</sub>, SO<sub>4</sub>, NH<sub>4</sub> in PM<sub>2.5</sub> or PM<sub>1</sub> | Aerosol Chemical Speciation Monitor (ACSM), Aerodyne Research Inc, USA |  |  |
| Black Carbon in PM<sub>2.5</sub> | Aethalometer AE33, Magee Scientific, USA |  |  |

### Respiratory function assessment

#### Spirometry

Our primary outcome, FEV_1_, will be assessed using a portable desktop computerized spirometer (Vitalograph 6000 Alpha Touch Spirometer), according to European Respiratory Society (Graham et al, 2019) and Association for Respiratory Technology and Physiology guidelines (Sylvester et al, 2020).

#### Fractional Exhaled Nitric Oxide

The fractional exhaled nitric oxide (FeNO) can be an indicator of airway inflammation. FeNO will be measured using NIOX VERO (Circassia AB) in accordance with manufacturer’s instructions and the American Thoracic Society guidelines (Khatri et al, 2021).

#### Oscillometry

Airway resistance (R_5_, R_20_ and R_5_-R_20_) and reactance (X_5_ and AX) will be measured using an oscillometer (Tremoflo® C-100) in accordance with manufacturer’s instructions and the European Respiratory Society guidelines (King et al, 2020).

#### Sample collection

##### Venepuncture

Blood samples will be obtained via venepuncture immediately before exposure and the day after exposure. A total of 10ml will be collected into EDTA coated Vacutainers at each sample timepoint. Samples will be centrifuged at 3000 RPM at 4°C for 15 min (Thermo Scientific Heraeus Multifuge 3 Plus Centrifuge). Plasma will then be aliquoted into clean polypropylene tubes and frozen at -80°C for later analysis and biobanking.

##### Nasal lavage

Nasal lavage samples will be collected using the nebuliser spray method (Mudway et al, 1999) at three time points: pre- and immediately post, and 24 hours post-exposure. Samples will be centrifuged to remove mucus and cellular components at 800xg at 4°C for 10 min (Thermo Scientific Heraeus Multifuge 3 Plus Centrifuge) and the cell-free supernatant aliquoted for longer-term storage at -80°C until required for analysis, or for longer-term storage as part of the sample biobank.

These samples will be used to test the hypothesis that exposure to NEE components causes changes in the immune status of the airway mucosa, skewing it toward a more Th2, Th17 profile consistent with an increased susceptibility of the airway to a range of asthma triggers. At the pre- and early post-exposure time point we will assess the concentration of a range of known Damage-Associated Molecular Pattern molecules (DAMPs) including High mobility group box 1 (HMGB1), the S100 Calcium Binding Protein A1 (S100A1), and the alarmins, interleukin-33 (IL-33), and Thymic stromal lymphopoietin (TSLP). In addition, we will also quantify the concentration of DNA complexes associated with myeloperoxidase (MPO-DNA), as a marker of netosis. At the pre and 24-hour post exposure time points a panel of Th1, TH2 and Th17 cytokines will be examined using BD Biosciences® Cytometric Bead Array (CBA) technology: IL-10, IL-6, IL-1β, IL12p70, IL-8, IL-4, IL-2, Tumour Necrosis Factor alpha (TNFα), nterferon-gamma (IFN-γ), Granulocyte-macrophage colony-stimulating factor (GM-CSF).

The lavage samples at the pre- and immediately post-exposure time points will also be analysed for a range of metals and metalloids to act as source-specific exposure biomarkers: Sb, Ba and Cu, (brake wear). Zn, (tyre wear), Fe, Mn, Mo (mechanical abrasion), Fe, Ca (road dust resuspension), and V, Ni and Cr as markers of diesel exhaust emissions. Analysis will be performed after nitric acid digestion by Inductively coupled plasma mass spectrometry (ICP-MS).

##### Urine samples

Urine samples will be collected by participants at home on the morning of site visits and during follow-up assessments the day after exposure. Urine samples will be aliquoted into clean 1ml polypropylene tubes and frozen at -80°C for later analysis and biobanking.

#### Pre exposure personal air pollution monitoring

To estimate any pre-testing exposure to PM, participants will be given a personal air quality monitor (Flow2, Plume Labs, Paris, France) to wear for three days prior to exposure visits to highlight any significant and possibly confounding exposure to high PM. Indications of very high exposure to PM prior to testing may be used to exclude participants from analysis. In addition, we will examine longer-term area level exposures based on PM_2.5_, PM_10_, NO_2_ and O_3_ concentrations measured at urban background monitoring stations in London. Background monitoring stations will be selected from sites within London’s Air Quality Network (https://www.londonair.org.uk/LondonAir/Default.aspx), based on their proximity to the residential address of the participants.

#### Exercise intensity

To account for changes in intensity during exercise, participants’ heart rate will be monitored for the duration of the exercise exposure using polar heart rate monitors (Polar H9). Data at minute five and minute 15 of each exercise bout will be averaged to assess for heart rate drift during the assessment. Borg scale Ratings of Perceived Exertion will be collected and assessed for the same reason.

#### Common allergy assessment

An additional 5ml blood sample will be collected to assess a full blood count including eosinophil levels, and specific immunoglobulin E levels across a panel of common aeroallergens to be included as covariates.

#### Symptoms surveys

Participants will complete an Asthma Control Test (ACT) (Nathan et al, 2004), an Asthma Quality of Life Questionnaire (AQLQ) (Khusial et al, 2020) and general symptom questionnaires at baseline and will be asked to complete a daily asthma questionnaire three days before and three days after each exposure (Martineau et al, 2015).

#### Air Pollution Measurements and Characterization of Non-exhaust Emissions

The air quality monitoring stations will provide measurement data according to the data quality assurance procedures described below, they will also enable a robust determination of the sources of exhaust, non-exhaust and other urban and regional sources of PM_10_ and PM_2.5_.

Mass concentrations of PM_10_ and PM_2.5_, oxides of nitrogen (NO, NOx, NO_2_) and ozone will all use reference measurement techniques supported by standardised operational and quality assurance approaches employed by the London Air Quality Network (www.londonair.org) or the UK government’s Automatic Urban and Rural Network (https://uk-air.defra.gov.uk/networks/network-info?view=aurn). Elemental composition will be measured at 1 hourly time resolution using ED-XRF (Tremper et al., 2018). To maximise size and chemical composition information, PM_2.5_ and PM_10_ will be collected on alternate hours using a switching valve and intermediate hours are interpolated as described by (Manousakas et al., 2022). Aerosol Chemical Speciation Monitor (ACSM, Ng et al., 2011) will measure the non-refractory composition of aerosol (Bressi et al., 2021) and black carbon will be measured using an Aethalometer.

The road surface conditions will be collected from surface sensors at Marylebone Road (DRS511, Vaisala, Finland) and using a camera at White City Flyover (DSC111, Vaisala, Finland), which will be used to differentiate wet and dry road surface conditions as per (Hicks et al., 2021).

#### Source Apportionment

The equipment described will identify sources of exhaust and non-exhaust concentrations by quantifying and source apportioning the high-time resolution chemical composition measurements described above to produce time series and factor profiles of source emissions in organic aerosols and elemental compositions. Using high-time resolution data in this way addresses many of the challenges faced when quantifying the different sources in the urban environment as the higher time resolution yields a greater variability from changing emission characteristics, boundary layer dynamics, and reflects short-lived events which are obscured by the long sampling time of filter-based approaches (Wang et al., 2018). This variability is fundamental to improving the performance of multivariate statistical approaches which are used to estimate the source contributions and fingerprints to solve a mass balance. Positive Matrix Factorization (PMF) (Paatero and Tapper, 1994) is the most commonly-used of these and the solutions describe the complex, time-dependent aerosol composition as a linear combination of factor profiles and their contributions. It has been used extensively for the identification of heavy metals, water soluble and carbonaceous sources (Hopke et al., 2020) and for high-time resolution elemental data from the Xact (Font et al., 2022b, Hasheminassab et al., 2020, Liu et al., 2019, Rai et al., 2020, Rai et al., 2021, Wang et al., 2018, Yu et al., 2019, Manousakas et al., 2022).

PMF will be applied to the PM_10_ and PM_2.5_ Xact data and ACSM data from all 3 stations using the Source Finder software (SoFi Pro, https://datalystica.com/) (Font et al., 2022b, Fröhlich et al., 2015, Manousakas et al., 2022, Reyes-Villegas et al., 2016, Visser et al., 2015b) to provide hourly source factor time series and factor profiles for the duration of the exposures. For the ACSM data, the SoFi software has been used in many recent studies to provide high-quality source information using a priori information (both source profiles and time series) and bootstrap resampling approach by following a standardized source apportionment protocol from Crippa et al. (2014) and Chen et al. (2022), which is now widely used in the source apportionment community (Via et al.,2021, Chen et al., 2021, Zhang et al., 2019, Tobler et al., 2020, Via et al., 2022). Recent work on data from Zurich (Manousakas et al., 2022) has shown that source apportionment of the XACT PM_10_ and PM_2.5_ data can be used to quantify the emissions from a range of sources in urban background locations.

Separating the tyre and road wear sources is not straight forward as they are internally mixed and generated by the same frictional forces. To help understand some of these processes in greater depth, additional size fractionated measurements of polymers are planned at Honor Oak Park and Marylebone Road during 2022/23 under a separate project (NERC, ‘Understanding UK airborne microplastic pollution: sources, pathways and fate’ - NE/T007605/1).

High volume PM_2.5_ samples (DHA-80, Digitel Elektronik AG, CH) will also be collected during the exposure periods and made available for additional laboratory analysis.

### Data analysis

#### Statistical methods

The main health outcome variables that will be considered as dependent variables are: FEV_1_ and FVC (differences between immediately, 24-hour and 3-day post-exposure and pre-exposure measurements); oscillometery, FeNO (same differences); asthma symptoms (yes/no) 24-hours and 3-days post-exposure; immune mediator concentrations HMGB1 and IL6 from nasal lavage (differences between 24-hour and 3-days post-exposure from pre-exposure values). The exposure covariates will be the integrated average concentration of the various pollutants and/or their sources during the exposure period. The results of the source apportionment will be used to evaluate the proportion of PM_2.5_ and PM_10_ mass related to each NEE and exhaust source. Additionally, PM_10_ and PM_2.5_ total mass will be used alternatively to check the main associations as a priori expected. Potential confounders will be included, such as meteorological variables including temperature, relative humidity, and gaseous pollutants during exposure. Adjustment for heart rate will additionally be added. Individual characteristics will also be considered, such as age, sex, severity of asthma, asthma control, quality of life, alcohol consumption, smoking and allergies.

Mixed effects regression modelling (as appropriate for the distribution of each health outcome variable) will be used to estimate the effects of different exposure variables on the range of acute response endpoints measured in the current study. A key timepoint for each outcome measure has been selected, based on previous observations in controlled exposure studies, or known temporal profiles of injury and inflammation (pre, 24-hour post) following exposures in inhaled toxicants (pre, post). Analysis will begin with single exposure covariate models for every source type, which will be used to build a multi-exposure model containing all source types. The effects of different exposures will be compared based on statistical significance and the effect size for selected exposure contrasts, such as a pre-defined change of concentration of the pollutant, or per interquartile range. A model will be applied contrasting the three sites (as categorical variables) to explore whether the profile of each site as a whole is important for the examined health effects. We will use R (version 4.1.3 or newer; R Development Core team) for the application of mixed models using functions from the lme4 and nlme packages.

Missing data will not be imputed. Likely causes of missing data are technical issues associated with health assessments in field environments. Due to the impact of technique on performance with spirometry in particular, missing data is likely. During their pre-testing visit, researchers will ensure that participants can complete all testing and sampling before they visit a testing site. The study sample size has been increased by 20% to accommodate missing data. Participants will also be given the opportunity to reschedule a visit if needed.

#### Ethics, safety, and dissemination

The study is Sponsored and managed by QMUL and supported by Imperial College London. This study has received HRA approval following favourable opinion from NHS REC (IRAS number: 320784). The study will be conducted according to the principles of the Helsinki Agreement (2013).

### Governance

An Independent Steering Committee (ISC) chaired by Prof Flemming Cassee will monitor and advise the study conduct and progress on behalf of the Sponsor and the HEI. The ISC will meet with the Chief Investigator and study team three times during the two-year study. At least one member of the of AUKCAR PPI group will be included in the meetings.

The study is Sponsored by QMUL (IRAS number: 320784) and is registered at clinicaltrials.gov. Monitoring and independent oversight of data is carried out by an independent assessor employed by QMUL and the audit team from the Health Effects Institute (https://www.healtheffects.org/).

### Consent

After discussing the study with the Principal Investigator, participants will have at least 24 hours to consider participation. To confirm consent, participants have the option of emailing signed consent forms to the Principal Investigator or signing consent forms at the first study visit.

### Data management

Survey Lab will be used to store electronic case report forms (CRFs). All data will be held on GDPR-compliant databases with integrated data validation checks and audit trails. All collected data will be held on backed-up encrypted servers. Any data transfers between ICL and QMUL will be performed via Secure File Transfer Protocols. Paper records, CRFs and consent forms and recruitment logs will be held locally in double locked locations at QMUL.

### Confidentiality

We will follow best practice guidelines provided in Standard Operating Procedures (SOPs) by the Pragmatic Clinical Trials Unit (PCTU) at QMUL. Paper records will be stored securely in locked filling cabinets in password locked rooms in the pass-protected Centre for Primary Care. Electronic records will be stored in a password-protected study database on a secure server, in the Centre for Primary Care. In the study database, personal details (name, address, date of birth) will be kept separate from research data, which will be identified by a unique study reference number. In tables of data, participants will only be identified by number, not by initials or name. Data management procedures will be completed in compliance with the GDPR and trial regulations. Survey reported data will be stored in the QMUL data safe haven, where data will be held in a UK server and access will be facilitated by two-factor authentication. Survey software with integrated data validation checks and audit trails will be used to record study data. Any data transfers between QMUL and Imperial College London will be completed via encrypted Secure File Transfer Protocol. All data will be backed-up weekly to ensure data is safeguarded from accidental loss. Data will only be accessed and used by those members of the research team at QMUL and Imperial College London and representatives of the Sponsor who have been granted permission.

### Record retention and archiving

In accordance with the UK Policy Framework for Health and Social Care Research, research records will be kept for 20 years after the study has been completed, while personal records will be stored for one year after study completion. At the completion of the study, data will be moved to a trusted archive centre. At the end of the retention period, data will be destroyed in accordance with best practice guidelines at the time of destruction.

Data Sharing: A copy of the study data will be held on the HDRUK BREATHE secure data hub https://www.breathedatahub.com. BREATHE’s mission is ‘Better respiratory health through better connected data’.

### Adverse events reporting

A risk and adverse events register will be maintained for the duration of the study. Accidents will be discussed with the Chief Investigator and reported promptly to the Sponsor as per Good Clinical Practice regulations and reviewed at study Project Management Group meetings.

### Dissemination plan and project outputs

Results arising from this study will be reported to the relevant funding agencies, scientific community and stakeholder groups through the following means:

1. Webinars on websites of our institutions, to provide more detailed summaries of results, with downloads of key documents.
2. Presentations, especially to London organisations including the GLA, councils and Health and Wellbeing Boards.
3. Peer reviewed publications.
4. Presentations at national and international conferences.

## Discussion

Internationally, policies to reduce pollution from traffic have predominately focused on exhaust emissions. This has been motivated by evidence of health effects associated with the proximity of populations to traffic sources and with proxies of primary exhaust emissions: black/elemental carbon, NO_2_ and PNC (HEI, 2010). However, as policy actions have driven reductions in exhaust emissions, attention has turned to NEE from vehicles arising from the wear of brakes, tyres and the road surface. These are currently unregulated and often exceed the total mass contribution of exhaust emissions in roadside and background PM_2.5_ and PM_10_. The significance of these sources will increase with greater penetration of non-internal combustion engines into the vehicle fleet as countries transition away from fossil fuels to meet their NetZero commitments. There is therefore an urgent need to establish the contribution of these sources to the adverse health effects attributed to ambient PM. Such information will be essential for the development of evidence-based policy to protect public health over the coming decades and to inform innovation within the automotive sector to reduce the risk of non-exhaust traffic emissions. While knowledge of the sources and compositional signatures of non-exhaust particles is becoming more established, significant uncertainty remains and there is limited information on their relative toxicity and health impacts.

Asthma is globally the most prevalent long-term condition, strongly impacted by air pollution, affecting adults and children, with major morbidity disproportionately affecting disadvantaged and minoritized ethnic populations. The asthmatic airway provides an exquisitely sensitive model to assess health impacts of non-exhaust emissions. The use of a quasi-experimental human real-world crossover design employed in this study allows the study of the health effects of non-exhaust emissions and to explore the hypothesis that clinically important adverse acute asthmatic responses are driven by non-exhaust components within coarse mode particulate matter.

Collectively this study will provide us with valuable information on the health effects of NEE components within ambient PM_2.5_ and PM_10_, whilst establishing a biological mechanism to help contextualise current epidemiological observations. Moreover, this work will establish key air quality and health variables to support larger epidemiological work exploring the health impacts of air non-exhaust air pollution, that will ultimately contribute to national air quality guidelines.

## Data Availability

All data produced in the present study are available upon reasonable request to the authors

## Contribution

JS, HH,MP,HW,DG,KK,IM,CG conceived the project. JS,MP,LM,IEB,HH,AT,HW,DG collected data.JS,HH,MP,AT,GC,DG,KK,IM,CG interpreted data. All authors contributed to the drafting of the paper.

